# Epidemiological profile and clinical conditions of women with or without precursorlesion for cervical cancer

**DOI:** 10.1101/2021.05.31.21258126

**Authors:** Lyvia Mirelle Carneiro de França, Caroline Ramalho Galvão, Lorena Ramalho Galvão, Maria Emília Cirqueira Silva, Bruna Matos Silva Dantas, Lorena Moura de Assis Sampaio, Magno Conceição das Mercês, Sarah dos Santos Conceição, Roberta Borges Silva, Julita Maria Freitas Coelho

## Abstract

**Introduction:** Cervical Cancer (CC) in Brazil is the fourth cause of death for malignant neoplasms in female population, being expected to 16,370 new cases for the biennium 2018-2019.

**Objective:** To identify factors associated with the presence of cytopathological changes of the cervix, HIV and/or syphilis in women who performed the Pap Smear (PS) at Family Health Unit (FHU) San Martin, in Salvador-BA, 09 March 2015 to 2018.

**Materials and methods:** This cross-sectional study consisted of 150 participants. After application of a questionnaire, there was conducted clinical examination of the breast and gynecological and rapid tests for diagnosis of syphilis and HIV types 1 and 2. The bivariate analysis was made by associating each one of the independent variables with the outcome, through contingency tables (Chi-square test and when necessary, the Fisher’s exact test). The estimates were obtained with Prevalence Ratio (PR) with their 95% confidence intervals (CI 95%) and p values. Mean differences were estimated through Student’s t-test. The goodness of the test was estimated by ratio of maximum likelihood (p ≤ 0.05).

**Results:** There were 11.33% of women with injury and 88.67% without injury. The group with injury (n = 17) presented the worst values in relation to the use of the cigarette (17.65% vs 3.76%) and clinical examination in breast change issues (17.65% vs 4.51%) and unloading (11.76% vs papillary 1.50%), in comparison to the group without precursor lesion pro CC, statistically significant (p ≤ 0.05). 5 cases were positive for syphilis (3.33%), and no case of HIV types 1 and 2.

**Discussion:** The study confirmed risk factors for injury precursors of CC, such as smoking and mammary changes, as well as how much research related to diverged condom use and reducing the risk of injury and the likely Association with Sexually Transmitted Infections (STI).

**Conclusion:** There was detected a low percentage of women with precursor lesions for the CC (11.33%), that were not associated with syphilis and HIV types 1 and 2, but were correlated the expected factors like mammary changes and smoking. The reorganization of actions of active search of women with risk factors for CC for the EP can culminate in the reduction of this cancer.

**Descriptors:** Pap Smear; Precursor Lesion; Cervical Cancer.

## INTRODUCTION

Cervical cancer (CC) is a health condition faced by women, and affects mainly developed countries. (XAVIER; TERRENGUI, 2006; AROSSI et al., 2003; VALE, et al., 2010). According to the world estimate of 570 thousand new cases, cervix cancer is on the fourth place of most frequent cases, representing 3.2% of all cancer types, and is also on the fourth place regarding mortality (WHO WHO, 2018). The 2018 estimate was 311,365 deaths worldwide. This corresponds to an estimated risk of 15,1 per thousand women (BRAY et al., 2018; FERLAY et al., 2018).

In Brazil, CC represented 7.4% of all cancer types, and was considered the fourth cause of death regarding malignant neoplasia in female population on 2018 (INCA, 2019).4The estimated incidence was 16,710 new cases of CC in the country for the triennium 2018-2019, which equates to an estimated risk of 15.43 per 100,000 women. In terms of mortality, in Brazil, in 2018, there were 6,526 deaths, and the crude mortality rate from cervical cancer was 6.10/100,000 (INCA, 2019). Other factors that increase the risk of developing this type of cancer are: early onset of sexual activity and multiple partners; smoking (the disease is directly related to the amount of cigarettes smoked); and prolonged use of birth control pills (INCA, 2019).

This disease is usually asymptomatic and has a slow-progression, and includes neoplastic or preneoplastic lesions, and Pap Smear (PS) can early detect it. CC screening measures or systematic screening for women are directed to performing the PS through effective, safe and low-cost public strategies (XAVIER; TERRENGUI, 2006; BAKER; MIDDLETON, 2003; SOLOMON; NAYAR, 2005; CAETANO et al, 2006; PINHO; FRANÇA JUNIOR, 2003).

In order to reduce morbidity and mortality from this neoplasm, since 1988, the Brazilian Ministry of Health (MS) follows as a norm the indication of the World Health Organization (WHO) to perform the cytopathological examination of the cervix every three years. .In addition, after two consecutive negative tests for neoplasms in women aged 25-63 years and/or women who have already had sexual activity (BRASIL, 2006; BRASIL, 2013). However, according to et a., cervical cancer control remains a challenge, causing improvements in prevention programs (TALLON et al., 2020).

The performance of PS in women with risk factors for CC is also an important strategy to reduce morbidity and mortality from this disease. Over the years several factors are being related to this pathology, such as Sexually Transmitted Infections (STIs); infectious and reactive conditions; sexual habits, such as early onset of early sexual life and many partners; active and passive smoking; and prolonged use of oral contraceptives (REZENDE, 1999; DIOGENES et al, 2001).

Thus, it is possible to envision a great professional relevance, at the service, academic and social, of studies about conditioning factors and determinants of cytopathological alterations that may have repercussions on CC, particularly for the early identification of women at higher risk and encourage the systematic performance of PS. Its results can support the elaboration and/or review of strategies for quality care, with excellence and humanized, as well as, can help in the reformulation of the work process of teams and/or health professionals.Considering the above context and in view of the prominent situation of cc in developing countries, this study aimed to identify factors associated with the presence of cytopathological changes of cervix, HIV and/or syphilis in women who performed the PS at the Family Health Unit (FHU) San Martin, in Salvador - BA, from 2015 to March 9, 2018.

## MATERIAL AND METHODS

We conducted a cross-sectional with women over 18 years of age who were users of the San Martin FHU in Salvador, Bahia, Brazil, whichwere subjected toPS between 2015 and March 9, 2018. This was a convenience sample which recruited women who agreed to participate in the study. This study was approved by the Research Ethics Committee of the State University of Feira de Santana, Bahia, Brazil, report N°. 2,548,705. All participants signed the Free and Informed Consent Form.

Women with a report presenting squamous and glandular cells and squamous report that presented precursor lesion for CC were eligible for the present study. Exclusion criteria were: women who presented results of PS with squamous report without precursor lesion for CC, indeterminate and or inconclusive; who no longer reside in the area covered by the unit during the period of data collection; who did not attend the unit for the interview on the scheduled date; and those that were not found in the residence during a home visit between 8 AM and 17 PM, during the period of data collection.

We obtained preliminary information through evaluation in medical records and a preventive collection record book, where women eligible for the study were identified. After this pre-selection, these women were located with the collaboration of Community Health Agents (CHA) or by telephone informed by them on the day of the examination collection. Through a home visit, the CHA invited them to participate in the research, and if accepted, a nursing consultation and interview was scheduled to be held at the FHU of the area, namely, FHU San Martin. We conducted the interview in an office from this FHU to obtain general, sociodemographic, life habits and health care data. In addition, we performed a nursing consultation in order to identify risk factors for CC, such as genital warts, from physical examination. We Measured weight using Welmy® anthropometric scale and height using a stadiometer, and body mass index (BMI) was calculated. Rapid tests were also performed for syphilis and HIV types 1 and 2, with advice before and after rapid tests, in order to assess possible individual risks and resolve any doubts of the participants.

The rapid test for HIV types 1 and 2 was HIV TRI LINE BIOCLIN, used to research HIV infection, based on immunochromatography technology. This test uses recombinant antigens HIV 1 and HIV 2, which react with antibodies present in serum, plasma and whole blood samples. The test for syphilis was the brand ALERE SYPHILIS, which presents a pre-coated membrane with recombinant antigen of *Treponema pallidum* in the T region. In order to perform them, a digital puncture with lancet was required, preceded by a careful washing and drying of the hands of the researcher and the participants, followed by antisepsis with 70% alcohol at the tip of the finger, an area that was used for puncture.

In the test for syphilis, 1 drop of blood and 4 drops of the reagent were placed in the device, and in the test for HIV types 1 and 2, 1 drop of blood in well A and 2 drops of the reagent in well Bwere placed in the device. Tests were validated through the appearance of line C. In cases not reactive for syphilis there appeared a line in the letter C, and in reagent cases there appeared two lines, one in C and one in area T. In the type 1 and 2 HIV test, in area T, we observed digits 1 and 2, which mark the test area and represent HIV-1 antigen and recombinant HIV-2 capture antigen. There was a negative testwhen there appeared only a the line in C, and positive when a line appeared in C and T, where in T1 it was considered positive for HIV 1 and T2 for HIV 2, and when presenting three lines the test was considered positive for both types of HIV. Both tests were performed from 5 to 30 minutes, where the result could later show a false positive. In case of positivity, we communicated the result oriented the woman regarding the relevant and appropriate measures. Data were also collected in medical records to complement clinical information, and the preventive collection record book was used to obtain the cytopathological examination report. Data analyses were performed with the aid of SPSS (Statistical Package for the Social Sciences) programs in version 22.0 (SPSS Inc., Chicago, IL, USA) and STATA version 11.0.

We obtained absolute and relative frequencies of all variables included in the study. For continuous variables, we obtained measures of central tendency (mean and their standard and median deviation). A bivariate analysis was carried out to identify the association of each of the independent variables with the outcome in question. The codependent variable was crossed, dichotomous, with each of the independent variables, according to its nature, by means of contingency tables (chi-square or fisher’s exact test). Prevalence Ratios (PR) were obtained with respective 95% Confidence Intervals (95% CI) and p value. Mean differences were estimated by t test. It was also verified the relative frequency of the types of cellular changes in PS detected in the sample.

## RESULTS

Between 2015 and March 9, 2018, 1,350 preventive studies were conducted at FHU San Martin. Among these, 392 reports (29.04%) were in accordance with the inclusion criteria of the study. However, there were only 52 present precursor lesions for CC (3.85%) and 45 had an unsatisfactory sample (3.3% of the tests). It is noteworthy that 330 tests could not be evaluated (24.44%), because the reports did not reach the FHU, even with the request for a second report, and/or did not have a detailed record in the exam control book.

The 392 women eligible for the study were invited to participate in the research through telephone calls, by invitations through the CHA and by home visits. In total, we identified 150 women that agreed to participate in the study, who would compose the comparison groups. Of the total women included, 17 (11.33%) presented precursor lesion for CC, while 133 (88.67%) were classified without injury (FIGURE 1). The mean age was 38.49 ± 13.68 years (median of 37 years), in an interval of 18 years to 81 years, and younger women predominated both with (70.59%) and without injury (54.89%). As for race/color, black and/or brown predominated in both groups (94.12% and 91.73% respectively). When investigating marital status, the highest percentage of women with precursor injury reported not having a partner (52.94%) versus 63.16% who had a partner among those without injury. Most of the sample had more than 7 years of schooling (58.82% vs. 63.91%) and did not work (52.94% vs 60.15%), in both groups (TABLE 1).

Regarding family income, most participants reported receiving up to one minimum wage (70.59% vs. 72.93%). Regarding family density, the majority reported living with up to two people at home (70.59% vs. 59.40%) and owning a home (58.82% vs. 58.65%). Regarding life habits, a statistically significant difference was observed only for the covariate smoking (p=0.016). However, we emphasize that the prevalence of smoking was higher in the group with injury (17.65%) than in the group without injury (3.76%). There were also low percentages of illicit drug users (5.88% vs. 3.01%). There was a predominance of women whopracticed physical activityup to once a week (82.35% vs. 78.95%) (TABLE 1).

**Table 1.** Sociodemographic characteristics and life habits of the sample according to the presence or absence of precursor lesions for CC. Salvador, BA, Brazil. (n 150)

| Chacateristics | Precursor Lesions for CC |  |  |  | *p* * |
| --- | --- | --- | --- | --- | --- |
|  | Yes (n=17) |  | No (n=133) |  |  |
| Age | N | % | n | % |  |
| ≤38.41 years | 12 | 70.59 | 73 | 54.89 | 0.219 |
| > 38.41 years | 5 | 29.41 | 60 | 45.11 |  |
| Mean±StandardDeviation: 38.49 ± 13.68 |  |  |  |  |  |
| Median: | 37 |  |  |  |  |
| MinimumandMaximum: | 18 - 81 |  |  |  |  |
| Race/skin color |  |  |  |  |  |
| White/Yellow/Indigenous | 1 | 5.88 | 11 | 8.27 | 0.733 |
| Black/Brown | 16 | 94.12 | 122 | 91.73 |  |
| Marital status |  |  |  |  |  |
| With a partner | 8 | 47.06 | 84 | 63.16 | 0.199 |
| Without a partner | 9 | 52.94 | 49 | 36.84 |  |
| Educationlevel |  |  |  |  |  |
| ≥ to 8 years | 10 | 58.82 | 85 | 63.91 | 0.682 |
| From zero to< 8 years | 7 | 41.18 | 48 | 36.09 |  |
| Occupation |  |  |  |  |  |
| Works | 8 | 47.06 | 53 | 39.85 | 0.569 |
| Does notwork | 9 | 52.94 | 80 | 60.15 |  |
| Family income** |  |  |  |  |  |
| ≤ 1 minimumwage | 12 | 70.59 | 97 | 72.93 | 0.838 |
| > 1 minimumwage | 5 | 29.41 | 36 | 27.07 |  |
| Household density(persons per household) |  |  |  |  |  |
| ≤ 2 persons | 10 | 58.82 | 78 | 58.65 | 0.989 |
| >2 persons | 7 | 41.18 | 55 | 41.35 |  |
| Own house |  |  |  |  |  |
| Yes | 12 | 70.59 | 79 | 59.40 | 0.374 |
| No | 5 | 29.41 | 54 | 40.60 |  |
| Smoker |  |  |  |  |  |
| No | 14 | 82.35 | 128 | 96.24 | 0.016 |
| Yes | 3 | 17.65 | 5 | 3.76 |  |

| Alcoholic beverage drinker |  |  |  |  |  |
| --- | --- | --- | --- | --- | --- |
| No | 7 | 41.18 | 64 | 48.12 | 0.589 |
| Yes | 10 | 12.66 | 69 | 87.34 |  |
| Illicit drug use |  |  |  |  |  |
| No | 16 | 94.12 | 129 | 96.99 | 0.534 |
| Yes | 1 | 5.88 | 4 | 3.01 |  |
| Physical activity |  |  |  |  |  |
| ≥ 2 days/week | 3 | 17.65 | 28 | 21.05 | 0.744 |
| 0 to 1 day/week | 14 | 82.35 | 105 | 78.95 |  |

\* Statistical significance level: $p \leq 0.05$
\*\* Minimum wage amount R\$954.00, on the date of data collection

**Figure 1.**
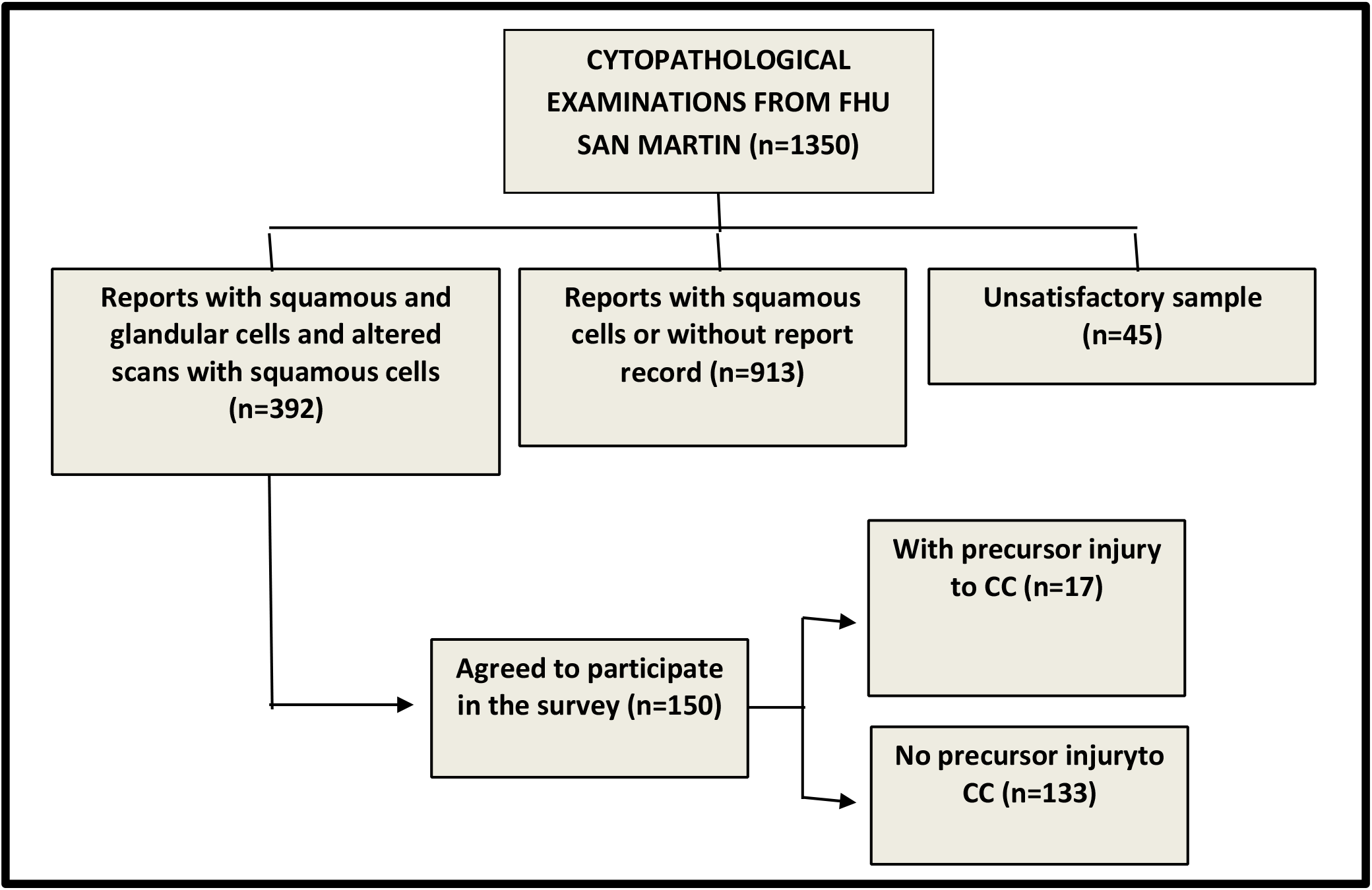
Flowchart of the selection and composition of the sample and comparison groups

Regarding the characteristics related to the general conditions of sexual health and life, it is noteworthy that, in general, the sample had inadequate weight in relation to height, since 82.35% of women with injury and 72.93% of those with no injury had BMI greater than 25. The first sexual contact was more frequent after 18 years (82.35% vs68.42%), and a greater number of women with number of partners ≤ 4 (52.94% vs69.92%) and also predominance of fixed partners (as) (94.12% vs 100%) (TABLE 2).When requesting information on gynecological characteristics, most women in both comparison groups reported having become pregnant more than once (88.24% vs. 89.47%), having undergone more than one delivery (88.24% vs 87.97%) and having more than one child (82.35% vs. 87.12%). It is worth noting that a percentage of women did not have an abortion was low (35.29% vs. 32.33%).

**Table 2.** Clinical, reproductive and family history of the sample according to the presence or absence of precursor lesions for CC. Salvador, BA, Brazil. (n =150)

| Characteristics | Precursor Lesions for CCC |  |  |  | *p* * |
| --- | --- | --- | --- | --- | --- |
|  | Yes (n=17) |  | No (n=133) |  |  |
| Weight | n | % | n | % |  |
| ≤ 71.69 | 7 | 41.18 | 75 | 56.39 | 0.235 |
| > 71.69 | 10 | 58.82 | 58 | 88.67 |  |
| Height |  |  |  |  |  |
| ≤ 1.58 | 8 | 47.06 | 61 | 45.86 | 0.926 |
| > 1.58 | 9 | 52.94 | 72 | 54.14 |  |
| BMI |  |  |  |  |  |
| ≤ 25 | 3 | 17.65 | 36 | 27.07 | 0.404 |
| > 25 | 14 | 82.35 | 97 | 72.93 |  |
| Amenorrhoea |  |  |  |  |  |
| No | 8 | 47.06 | 70 | 52.63 | 0.665 |
| Yes | 9 | 52.94 | 63 | 47.37 |  |
| First sexual contact |  |  |  |  |  |
| ≤ 18 years | 3 | 17.65 | 42 | 31.58 | 0.238 |
| > 18years | 14 | 82.35 | 91 | 68.42 |  |
| Number of sexual partners |  |  |  |  |  |
| ≤ 4 partners | 9 | 52.94 | 93 | 69.92 | 0.157 |
| > 4 partners | 8 | 47.06 | 40 | 30.08 |  |
| Fixed sexual partner |  |  |  |  |  |
| Yes | 16 | 94.12 | 133 | 100.00 | 0.005 |
| No | 1 | 5.88 | 0 | 0 |  |
| Pregnancies |  |  |  |  |  |
| None | 2 | 11.76 | 14 | 10.53 | 0.876 |
| 1 or more | 15 | 88.24 | 119 | 89.47 |  |
| Delivery |  |  |  |  |  |
| None | 2 | 11.76 | 16 | 12.03 | 0.975 |
| 1 or more | 15 | 88.24 | 117 | 87.97 |  |
| Offspring |  |  |  |  |  |
| None | 3 | 17.65 | 17 | 12.88 | 0.587 |
| 1 or more | 14 | 82.35 | 115 | 87.12 |  |
| Misscarriage |  |  |  |  |  |
| None | 11 | 64.71 | 90 | 67.67 | 0.806 |
| 1 or more | 6 | 35.29 | 43 | 32.33 |  |
| Interval between PS |  |  |  |  |  |
| Less than 1 year | 4 | 23.53 | 16 | 12.03 | 0.189 |
| 1 year or more | 13 | 76.47 | 117 | 87.97 |  |
| Breastfeeding |  |  |  |  |  |
| Yes | 13 | 76.47 | 109 | 81.95 | 0.585 |
| No | 4 | 23.53 | 24 | 18.05 |  |
| Contraceptive methods |  |  |  |  |  |
| No | 8 | 47.06 | 44 | 33.08 | 0.254 |
| Yes | 9 | 52.94 | 89 | 66.92 |  |
| Use of hormonal contraceptives |  |  |  |  |  |
| No | 9 | 52.94 | 75 | 56.39 | 0.787 |
| Yes | 8 | 47.06 | 58 | 43.61 |  |
| Correct use of hormonal contraceptive |  |  |  |  |  |
| No | 15 | 88.24 | 120 | 90.23 | 0.797 |
| Yes | 2 | 11.76 | 13 | 9.77 |  |
| Condom use |  |  |  |  |  |
| Yes | 1 | 5.88 | 16 | 7.52 | 0.807 |
| No | 16 | 94.12 | 123 | 92.48 |  |
| Correct use of condom |  |  |  |  |  |
| Yes | 1 | 5.88 | 8 | 6.02 | 0.983 |
| No | 16 | 94.12 | 125 | 93.98 |  |
| Hypertension |  |  |  |  |  |
| No | 14 | 82.35 | 101 | 75.94 | 0.556 |
| Yes | 3 | 17.65 | 32 | 24.06 |  |
| Diabetes |  |  |  |  |  |
| No | 15 | 88.24 | 124 | 93.23 | 0.457 |
| Yes | 2 | 11.76 | 9 | 6.77 |  |
| Family history of breast cancer |  |  |  |  |  |
| No | 15 | 88.24 | 111 | 83.46 | 0.613 |
| Yes | 2 | 11.76 | 22 | 16.54 |  |
| Family History of CC |  |  |  |  |  |
| No | 16 | 94.12 | 121 | 90.98 | 0.665 |
| Yes | 1 | 5.88 | 12 | 9.02 |  |
| Syphilis |  |  |  |  |  |
| No | 16 | 94.12 | 129 | 96.99 | 0.534 |
| Yes | 1 | 5.88 | 4 | 3.01 |  |

\* Statistical significance level: $p \leq 0.05$

Regarding the interval between the last two preventive tests, the groups also indicated that this interval was above one year (76.47% vs. 87.97%) (TABLE 2).Most women did not use hormonal contraceptive methods (52.94% vs. 56.39%) and a high percentage did not use male or female condoms (94.12% vs. 92.48%). It is worth noting that the women who participated in the study used only one contraceptive method. Regarding the testing of HIV types 1 and 2, no positive women were identified for this disease. Regarding syphilis, 05 women showed a positive result (5.88% with lesions and 3.01% without injuries). In the groups exposed or not exposed to cc precursor lesions, it was observed that there was no statistically significant difference for these variables between the comparison groups, demonstrating homogeneity between them (TABLE 2).

When analyzing the mammarian conditions, vaginal microflora and other gynecological characteristics, it was noticed that mammarian palpation (p=0.032) and papilar discharge (p=0.013) were different from the statistical point of view between the groups in comparison. However, the prevalence of alteration in mamamarian palpation was higher in the group with injury (17.65%) than in the group without injury (4.51%). Similarly, the presence of papilar discharge was higher in the group with injury compared to the opposite group (11.76% vs 1.50%) (TABLE 3).

**Table 3.** Mammarian conditions, vaginal microflora and other gynecological characteristics presented by the sample according to the presence or absence of precursor lesions for CC. Salvador, BA, Brazil. (n =150)

| Characteristics | Precursor Lesions for CC |  |  |  | *p* * |
| --- | --- | --- | --- | --- | --- |
|  | Yes (n=17) |  | No (n=133) |  |  |
| Breasts | n | % | n | % |  |
| *Symmetric* | 16 | 94.12 | 110 | 82.71 | 0.227 |
| *Asymmetric* | 1 | 5.88 | 23 | 17.29 |  |
| Mamarian palpation |  |  |  |  |  |
| *Normal* | 14 | 82.35 | 127 | 95.49 | 0.032 |
| *Altered* | 3 | 17.65 | 6 | 4.51 |  |
| Papilar discharge |  |  |  |  |  |
| *No* | 15 | 88.24 | 131 | 98.50 | 0.013 |
| *Yes* | 2 | 11.76 | 2 | 1.50 |  |
| Presence of vulvar lesion |  |  |  |  |  |
| No | 16 | 94.12 | 128 | 96.24 | 0.674 |
| Yes | 1 | 5.88 | 5 | 3.76 |  |
| Presence of vaginal injury |  |  |  |  |  |
| No | 17 | 100 | 132 | 99.25 | 0.720 |
| Yes | 0 | 0 | 1 | 0.75 |  |
| Leucorrea |  |  |  |  |  |
| No | 8 | 47.06 | 59 | 44.36 | 0.833 |
| Yes | 9 | 52.94 | 74 | 55.64 |  |
| Microflora |  |  |  |  |  |
| Physiological | 11 | 64.71 | 93 | 69.92 | 0.660 |
| Pathogenic | 6 | 35.29 | 40 | 30.08 |  |

\* Statistical significance level: $p \leq 0.05$

Regarding the presence of precursor lesions for CC, 11.33% of the women interviewed presented this condition (n=17). Regarding the distribution of these aforementioned lesions, a frequency of 4.67% of atypia of indeterminate significance was observed in possibly non-neoplastic squamous cells (ASCUS); 3.33% of low-grade intraepithelial injury (including HPV, mild dysplasias and CIN I) (LSIL); 1.33% of atypias of indeterminate significance in possibly non-neoplastic squamous cells, and cannot rule out high-grade intraepithelial injury (ASC-H); 1.33% of high-grade intraepithelial lesion (including CIN II and III, carcinoma in situ) (HSIL); 0.67% of atypias of indeterminate significance in glandular cells (GCA), possibly non-neoplastic, and cannot rule out high-grade intraepithelial lesion (GRAPH 1).

**Graph1.**
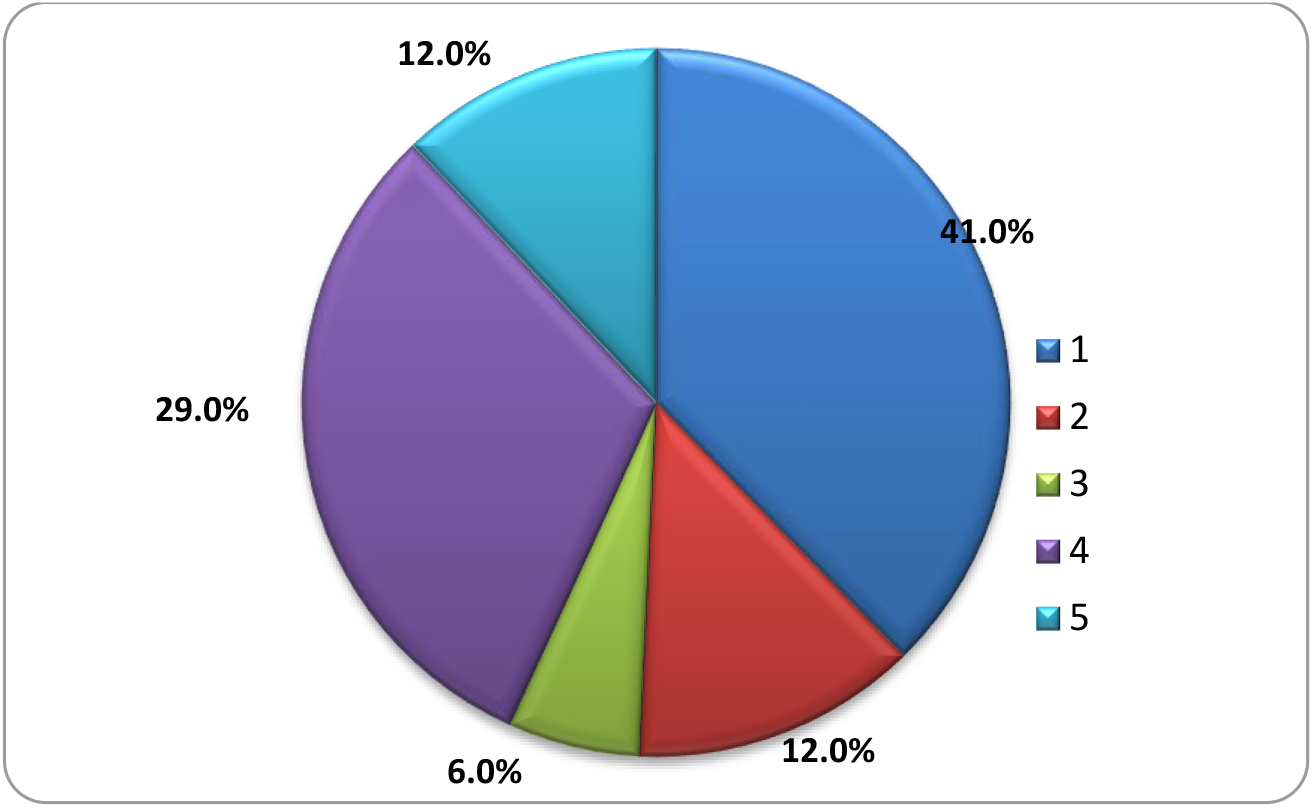
Distribution of types and percentages of precursor lesions for possibly non-neoplastic cervical cancer detected in Pap smear tests in women using the San Martin FHU (n=17). Salvador, 2018.

## DISCUSSION

The present study detected a relatively low percentage of CC precursor lesions, no HIV-type and or type 2 positive cases, and 5 cases of syphilis by rapid testing. It is important to highlight that the screening policy for CC considers a threshold of 1% for unsatisfactory samples, as a factor that evaluates the quality of the collection and conservation of the PS sample. It is noteworthy that in Ceará, Paraíba, Pernambuco, Alagoas and Bahia this coverage reached rates higher than 1% for tests with unsatisfactory results (SILVA, 2011), a finding confirmed in this study where 3.33% of the reports are unsatisfactory. A low percentage of 29.04% tests with ectocervix cells (squamous and stratified epithelium) and endocervix cells (simple columnary epithelium) and endocervix cells (simple columnary epithelium) were also detected, against a higher percentage (43.19%) of slides collected without the presence of endocervical cells, and such concomitant presence is considered as a relevant characteristic for the evaluation of the quality of the test. A study conducted at the Office of Integral Assistance to Women’s Health of Tiradentes university of Aracaju verified a greater number of tests with squamous and glandular cells 46.2%, against 45.1% of squamous cells, the latter with results very close to that found in the present (FERREIRA et al, 2015). Therefore, the quality of these tests included in the studies cited can be considered good.

In general, the mean age of women who underwent the test (38.41) was lower. A lower percentage was obtained in a study conducted in Campinas-SP, which verified amongst 54,33 cytopathological tests performed, 24.6% of women were under 25 years of age, 68.8% between 25 and 59 years and 6.5% aged 60 years or older (FREITAS et al, 2006). This may explain the higher frequencies of some non-health-friendly habits in the women investigated, such as condom use and regularity of PS. Regarding race/color, most of the women in the sample were black or brown (with 94.12% injury and without injury 91.73%) and with reported up to four sexual partners (with 52.94% injury and without injury 69.92%). A study in the North region revealed a similar study, where the majority of women 89.1% were brown and 70% had more than two sexual partners throughout their lives (PRADO et al, 2012). Hence, it is possible to suspect the influence of race/color on the higher risk for syphilis, and STIs in general, as well as for the occurrence of CC lesions.

In the present investigation, it was also observed that most women with eight or more years of schooling (with injury 58.82% and without injury 63.91%), with a partner (with injury 47.06% and without injury 63.16%) and family income up to one minimum wage (with injury 70.59% and without injury 72.93%). In another study conducted in Jequié-BA, a similar result was found, since 58.6% of the women who underwent the PS had a partner and 77.1% had an income of up to 01 minimum wage (SILVA, 2014). However, in the schooling item there was a discrepancy in relation to this same study, since a higher percentage of women positive for CC lesions with higher schooling was found. The importance of greater education in women’s treatment with preventive performance is highlighted, thus, the higher the level of education, the better the level of understanding about the CC, its risk factors, diagnosis and treatment (CORREA, 2009). There is the possibility of a positive effect on the level of education and the low frequency of syphilis and other unfavorable conditions, such as the presence of lesions for CC in the women investigated here. Regarding lifestyle, in the present sample, women who practice 1 day of physical activity or do not practice it are between 78.95% (without injury) and 82.35% (with injury). In a study in Rio Branco, there was a higher prevalence of preventive examination in women who practiced physical activity (82.2%) (BORGES et al, 2012). Regarding smoking and alcohol consumption, the results showed that most women were not considered smokers or alcoholics. This is in accordance with what was observed in a research developed in Bebedouro-SP with the old women who performed the PS, it was found that most participants also did not consume cigarettes or alcoholic beverages (OLHE et al, 2013). This low frequency is a positive characteristic in the present sample, considering the greater vulnerability of women to the effects of tobacco and alcohol on health as a whole.

When we investigated characteristics related to the presence of precursor lesion for CC, it was observed that 82.35% started sexual life aged 18 years, 94.12% had a fixed partner, 82.35% had children, and 52.94% did not use hormonal contraceptive method. It is emphasized that according to Ferreira et al (2015), the contraceptive methods most used by women who underwent PS, were hormonal contraceptive (39%), condom (14.7%), tubal ligation (1.8%), adhesive (0.6%) and vaginal ring (0.3%). However, in a research developed at the Centro Viva Vida in Minas Gerais, a divergent result was verified from the ones obtained regarding the presence of cc precursor lesion, since 57% of the women studied who presented precursor lesions for CC started sexual life between 15 and 19 years. Regarding the use of hormonal contraceptive methods, Figueiredo et al. detected similar rates since only 46.2% of their sample reported this use, 67.7% did not report having multiple partners, and 58,1% had between 2 and 4 children (FIGUEIREDO et al, 2015). Thus, with regard to the rates of the present study mentioned above, it can be seen that the women investigated may be considered at lower risk for the occurrence of STIs and precursor lesions of CC. However, they suggest a greater reflection on the need for greater encouragement and information on the part of women in order to increase the frequency of safer sexual habits.

Regarding the interval between preventive cervical cancer tests, there was a high percentage of women who underwent PS over one year, both among those who presented a precursor lesion of CC (70.59%) as well as those that did not show such a condition 81.95%. In a study on the knowledge and attitudes of women about the cytopathological examination of CC, Silva (2015) found that 57.9% of the women in the study performed the PS annually, 41% above one year, and 1.1% had never performed this test. Whereas cellular changes that can evolve into CC can be easily discovered by PS, and consequently have high chances of cure in general (INCA, 2019), these findings suggest a success in health actions focused on women’s health in the aforementioned area of coverage of the FHU investigated.

However, the present study detected a low population’s support for female or male condom use, considering that only 5.88% of women with injury and 7.52% of women without injuries reported this use. This is in accordance with the results of other studies that also investigated these characteristics, reaffirming the challenge for further expansion of this practice (SILVA et al, 2017; FERREIRA et al, 2015; BOLDRINI, 2012). It is worth adding that in the sample each participant used only one contraceptive method, emphasizing that the majority’s concern was non-conception. Therefore, actions aimed at encouraging greater protection in the sexual life of these women are necessary, and not only the possibility of pregnancy. Especially in the greater clarification about the possibility of contracting STIs in the face of non-use of condoms, especially syphilis and HIV.

Regarding the type of vaginal secretion in PS, 44.67% of women presented physiological secretion. Although the sample shows large percentages of *Lactobacillussp*, coccus and other bacilli in microbiology (64.71% with lesion vs 69.92% without injury), this result is considered normal because they are part of the vaginal microbiota, not characterizing infection. Although this sample presented large percentages of Lactobacillussp, coconuts and other bacilli in microbiology (64.71% with lesion vs 69.92% without injury), this result is considered normal because they are part of the vaginal microbiota, not characterizing infection. However, Leal et al (2003), analyzing the results of gynecological tests and the microbiology of the Oncology Control Center of Acre, evidenced the presence of normal vaginal secretion in 93.4% (n=2,239). This lower rate of physiological secretion is sometimes detected because it was derived from the predominance in the sample of women with 4 or more partners, as well as the frequency of interval between preventive tests greater than one year. Thus, in relation to these variables, these women lack greater information and greater stimulation of healthier sexual practices.

On the other hand, it is also important to note that only 35.29% of the sample with precursor lesions for CC presented microflora considered pathogenic. Under study Silva18 with 415 women, 86.8% of them had normal preventive results, which is close to these results that showed that 133 women had results considered within normal limits (88.67%). A low percentage of women with lesions was also detected, 4.67% of which were ASCUS; 3.33% LSIL; 1.33% ASC-H; 1.33% HSIL; and 0.67% of GcA. Data found in research in the Northeast region also showed low percentages of lesions, and reported similar values for LSIL (1.2%) and HSIL (0.30%)(OLIVEIRA; ALMEIDA, 2013). These results may represent good gynecological health in this sample. Certainly these findings must have influenced the low frequency of syphilis and absence of HIV in the women investigated. That is, they point to low risk in relation to such characteristics.

Regarding the frequency of syphilis by rapid testing in the sample, a small proportion of positive women were found (5.88% with lesion vs. 3.01% without precursor lesions for CC). In 2016 alone, a total of 87,593 cases of syphilis acquired in Brazil were reported, and of these 3,508 were in Bahia, currently considered a region with syphilis epidemic (BRASIL, 2017). Thus, it is possible that the population of this area has really low rates, or the low power of the sample did not allow estimating the real prevalence. Therefore, it is suggested a thorough investigation of the possible partners of these positive women, through survey of information with them and followed by an active search of these individuals, in order to try to interrupt a possible chain of transmission in this group.

Finally, considering the conditions for its performance, this research allowed the obtaining of the profile of the women who composed the sample, in the presence or absence of precursor lesions for the CC. The potential benefit of this information in the review of strategies for prevention, detection and coping with syphilis by the FHU to which they belonged is highlighted. However, the present investigation presented limitations such as the reduced size of the sample employed, which did not allow enough power to detect real differences between the groups investigated.

In addition, the time of study conduction was limited, since it was carried out as a master’s activity, a course that has its own academic norms for performing the stages and maximum completion period defined for research that will generate the student’s final dissertation. Moreover, it did not have external funding that would allow better conditions for its realization, such as aggregating a team of interviewers and/or nurse examiners, in order to speed up the performance of data collection. Only one interviewer, who also performed the process of obtaining consent to participate in the research, conducted all interviews, rapid tests and counseling to the participants, as well as made the appropriate referrals for treatment when they were necessary. Another point to highlight, it refers to the convenience sample, with a predefined population and limited to women users of health services of the FHU San Martin, in the municipality of Salvador, Bahia, which limits the generalization capacity of the study. This may have made it difficult to obtain more representative rates of this population, as well as limitations for possible statistical inferences.

However, since the literature refers to STIs as a risk factor for CC, most studies in this line are more targeted for more specific detection of the presence of HPV virus than HIV 1 and 2 and *Treponema pallidum* bacteria in populations. In the present investigation, it sought several factors that could be related to the presence of precursor lesions of cervical cancer, considering the magnitude of the rates of this pathology, as well as its effect on the reduction and potential years and quality of life of women. Thus, investigations of this nature are relevant in order to expand the body of evidence on the theme. Finally, given the non-detection of positive cases of HIV 1 and 2 represented a positive aspect from the epidemiological point of view of STIs, this made it impossible to study factors related to this disease. Thus, it would be necessary to expand the present to clarify the real frequency rate of women with HIV 1 and 2 in this area of coverage.

## CONCLUSION

These results may be very useful for reviewing strategies aimed at identifying and preventing conditioning factors and determining cytopathological changes and their effects on the occurrence of CC. In particular, they may raise a greater interest in the stimulation and early uptake of women with such conditions and/or to perform PS. Thus, they will be able to support the development of new strategies for quality care, with excellence and humanized, and assist in the reformulation of the work process of this team. In general, it is expected that the information obtained in this investigation stimulates interest in further studies of this nature, given the burden that CC and syphilis can bring to women, especially those at higher risk, such as congenital syphilis in their children, reduction of potential years of life, and individual and public financial costs.

This research can be of great professional relevance and to the service investigated and similar, in addition to its academic and social importance. In relation to professionals and the service, the results may contribute to a rethinking about the need to identify conditioning factors and determinants of cytopathological changes, especially those that can facilitate the occurrence of STIs, especially with the recent increase in syphilis rates in Brazil.Likewise, they can add subsidies in the reformulation of the work process of this team, and redirect it to groups more vulnerable to such injuries. On the other hand, they can have a positive impact on the choice of more specific actions to capture positive women early and to increase the performance of PS, which are necessary, with a view to renewing strategies for quality care, with excellence and humanized.

At the academic and social level, this study, the scientific knowledge obtained on the subject may be to help in the planning of future research aimed at the screening and estimation of risk factors for the occurrence of the aforementioned diseases. In addition, the use of testing for HIV types 1 and 2 and syphilis allowed the early identification of positive cases of syphilis, and revealed the absence of hiv-positive women 1 and 2. Thus, these women had the chance to be referred/oriented to immediate treatment, besides being properly informed about such injuries. Similarly, women positive for lesions of CC precursors, and other injuries such as the presence of palpable breast masses, papilar discharge and cytopathological alterations in PS, could be identified and properly clarified about the need to treat such conditions in order to reduce the risk for the development of breast and/or cervical cancer. Finally, five cases of previously unknown syphilis could be detected in the women investigated. This indicates the presence of this disease in this area of coverage, considering the warning for an increase in its frequency, and its possible serious consequences such as congenital syphilis that can cause blindness in the fetus.

In summary, more studies in this line of research are suggested that may reinforce a more assertive prevention of gynecological diseases, HIV 1 and 2, syphilis and other STIs, especially those with larger samples that can estimate the real effect of the risk factors studied and directed to subgroups in greater vulnerability to such outcomes.

## Data Availability

The authors declare availability to present data mentioned in the manuscript.

## ACKNOWLEDGEMENTS

To the State University of Feira de Santana, Bahia, Brazil, and especially the Center for Research, Integrated Practice and Multidisciplinary Research (NUPPIIM) for the support for this research, and professionals and users of the FHU San Martin, as well as the municipality of Salvador-BA, which not only allowed its realization, but also collaborated with the availability of adequate physical space for data collection and kits for rapid testing tests for syphilis and HIV Bahia, Brazil for its contribution to research.

